# Loneliness and Cognitive Decline Among U.S. Adults: A Stratified Analysis of the BRFSS

**DOI:** 10.64898/2025.12.10.25342005

**Authors:** Mojisola Fasokun, Temitope Ogundare, Fadeke Ogunyankin, Kaelyn Gordon, Seun Ikugbayigbe, Mariam Michael, Kakra Hughes, Oluwasegun Akinyemi

## Abstract

**Background:** Loneliness is an emerging public health concern linked to adverse mental and physical outcomes. It may play a key role in cognitive aging, yet its population-level association with subjective cognitive decline (SCD) across demographic groups is not well characterized. We evaluated how the frequency of loneliness relates to SCD in U.S. adults and whether associations differ by sex, age and race/ethnicity.

**Methods:** We performed a cross-sectional analysis of adults aged ≥16 years using nationally representative 2016–2023 Behavioral Risk Factor Surveillance System data (BFRSS). Loneliness was categorized as never, rarely, sometimes, usually or always. The primary outcome was self-reported SCD in the past year. Survey-weighted logistic regression models adjusted for sociodemographic factors, health insurance, metropolitan status and survey year were used to estimate adjusted marginal probabilities of SCD across loneliness categories. Interaction terms and stratified margins evaluated effect modification by sex, age group (16–44, 45–64 and ≥65 years) and race/ethnicity (non-Hispanic White, non-Hispanic Black and Hispanic).

**Results:** Among 85,969 adults who reported loneliness, 13,879 (16.2%) experienced subjective cognitive decline (SCD), with a mean age of 65.7 ± 10.6 years. Loneliness showed a strong dose–response relationship with SCD. Predicted probabilities of SCD increased from 9.9 % (95 % CI, 9.3–10.5 %) among respondents who never felt lonely to 15.0 % (14.1–15.9 %) for rarely, 24.9 % (23.6–26.1 %) for sometimes, 38.4 % (34.4–42.5 %) for usually and 45.7 % (41.0–50.4 %) for always lonely adults (p < 0.001). Women who were always lonely had an adjusted probability of SCD that was 10.7 percentage points higher than men; sex differences were negligible at lower loneliness levels. Age differences were minimal across most loneliness categories; however, among adults who were always lonely, those aged >64 years had significantly lower predicted cognitive function compared with adults aged 18–64 years (p < 0.001). Racial and ethnic differences were modest; the only significant contrast was a 1.7 percentage-point lower probability of SCD for non-Hispanic Black adults compared with Whites among those who never felt lonely. Other subgroup differences were not statistically significant.

**Conclusions:** Loneliness is independently and strongly associated with higher likelihood of subjective cognitive decline among U.S. adults, and this relationship is most pronounced for chronic loneliness. While sex and age modified the effect of loneliness, racial/ethnic disparities were minimal. These findings identify loneliness as a modifiable social determinant of cognitive health, supporting the need for broad social connection initiatives and targeted efforts for women and mid-life adults with chronic loneliness.

## Introduction

Loneliness is increasingly recognized as a serious public health issue with wide-ranging implications for mental, physical and cognitive health(1–3). Defined as a distressing subjective state arising when perceived social connections are inadequate(3, 4). Loneliness affects a substantial proportion of adults in the United States: about one third of adults aged 45 and older experience loneliness, and nearly one quarter of those aged 65 and older are socially isolated(5, 6). Loneliness and social isolation are related but distinct constructs(2, 7, 8), and the present study focuses on loneliness. Feelings of loneliness have become so pervasive that some researchers describe them as a “loneliness epidemic”. Beyond its psychosocial toll, loneliness has been linked to increased mortality risk, cardiometabolic disease, depression, and cognitive decline(9, 10). Understanding and addressing loneliness is therefore a pressing public health priority.

Cognitive decline and dementia represent another growing challenge as populations age(11–13). More than 55 million people currently live with dementia worldwide, a figure projected to rise to 78 million by 2030(14, 15). In the United States, about 10–11% of adults aged 45 and older report subjective cognitive decline, corresponding to roughly 1 in 9 people in this age group(16). While age is the strongest risk factor, a number of potentially modifiable factors may influence cognitive trajectories(13, 17, 18). Emerging evidence suggests that loneliness predicts poorer cognitive performance and faster rates of decline across multiple domains(19–21). Proposed mechanisms include chronic activation of stress pathways leading to elevated cortisol and inflammation, reductions in cognitive stimulation, and depressive symptoms that mediate the loneliness–cognition link(22–24). Yet many questions remain. Much of the existing literature has relied on small, localized or clinical samples of older adults(25, 26). Studies have often been cross-sectional, limiting inference about directionality, and have rarely explored how the loneliness–cognitive decline relationship varies by sex, race/ethnicity or age group(19, 27, 28). Furthermore, loneliness is measured inconsistently across studies, complicating synthesis of results.

To address these gaps, we analyzed nationally representative data from the 2016–2023 Behavioral Risk Factor Surveillance System (BRFSS)(3) to examine the association between self-reported loneliness and subjective cognitive decline (SCD) in U.S. adults. The BRFSS cognitive module asks respondents whether, during the past 12 months, they experienced confusion or memory loss that is worsening; a measure that has been linked to subsequent objective cognitive impairment and dementia risk(29). Our study makes several contributions. First, we evaluate a dose–response relationship by leveraging the BRFSS loneliness question, which categorizes the frequency of loneliness (never, rarely, sometimes, usually, always), rather than using a binary measure. Second, we apply complex survey weighting and inverse probability weighting to approximate causal effects of loneliness on cognitive decline while controlling for sociodemographic and structural factors. Third, we explore whether associations differ by sex, age, and race/ethnicity, groups for whom experiences of loneliness and cognitive aging may vary. By analyzing a large, diverse sample with rigorous methods, this study aims to clarify the role of loneliness as a potentially modifiable risk factor for cognitive health across the life course. We hypothesized that higher levels of loneliness will be associated with greater likelihood of subjective cognitive decline and that these associations may differ by demographic subgroup.

## Methodology

### Study design and data source

This cross-sectional study analyzed data from the Behavioral Risk Factor Surveillance System (BRFSS) collected between 2016 and 2023(30). The BRFSS is an ongoing, nationally representative health survey conducted annually by the U.S. Centers for Disease Control and Prevention (CDC)(31). It employs a complex, multistage sampling design with stratification, clustering, and unequal probabilities of selection. Interviews are conducted via landline and cellular telephones across all 50 U.S. states, the District of Columbia, and U.S. territories. Survey weights provided by the CDC account for sampling design, non-response, and post-stratification, enabling generalization to the non-institutionalized adult population(3). Because the BRFSS uses de-identified, publicly available data, institutional review board approval was not required; the study adhered to ethical principles outlined in the Declaration of Helsinki.

### Study population

We included BRFSS respondents aged 18 years or older who participated in surveys between 2016 and 2023 and had complete data on loneliness, subjective cognitive decline (SCD), and covariates. Respondents were excluded if they responded “don’t know/not sure,” “refused,” or had missing responses for any key variable. To ensure comparability, we also excluded individuals with missing state or year identifiers, which were used as fixed effects. After applying these criteria, the final analytic sample comprised 86,520 participants. Survey weights were applied in all analyses to account for the probability of selection and to generate nationally representative estimates.

### Exposure: loneliness

Loneliness was assessed using the BRFSS question: “How often do you feel lonely?” with five response categories: “Always,” “Usually,” “Sometimes,” “Rarely,” and “Never.” We treated loneliness as an ordinal categorical variable, with increasing levels representing greater perceived social isolation. This gradation allowed exploration of dose–response relationships between loneliness and cognitive decline.

### Outcome: subjective cognitive decline

Subjective cognitive decline was measured using the BRFSS module question: “During the past 12 months, have you experienced confusion or memory loss that is happening more often or getting worse?” Responses were coded as yes or no. Although self-reported, this measure has been used widely as an early indicator of cognitive impairment and is associated with objective cognitive performance and dementia risk(32–34).

### Covariates

Covariates were selected based on prior research linking sociodemographic factors to loneliness and cognitive health. They included: age (continuous), sex (male or female), race/ethnicity (non-Hispanic White, non-Hispanic Black, Hispanic, Other), education (less than high school, high school graduate, some college, college graduate), marital status (married, divorced/separated, never married, widowed), employment status (employed, unemployed, retired, unable to work), health insurance type (private, Medicare, Medicaid, self-pay, other), metropolitan status (metropolitan vs. non-metropolitan), urbanicity (urban vs. rural), and language spoken at home (English, Spanish, Other). State of residence and survey year were included as fixed effects to control for geographic and temporal heterogeneity. All categorical covariates were dummy-coded for inclusion in regression models.

### Statistical analysis

We estimated associations between loneliness and subjective cognitive decline using survey-weighted logistic regression models. The dependent variable was SCD (1 = yes, 0 = no), and the main exposure was the loneliness category (reference group = “Never”). Models were adjusted for all covariates listed above and incorporated BRFSS sampling weights and design variables (primary sampling unit and strata) via the Stata svyset command. To account for complex survey design, we used Taylor-series linearization to obtain robust standard errors clustered at the primary sampling unit level.

To aid interpretation, we calculated adjusted marginal probabilities of SCD for each loneliness category using Stata’s margins command. These margins represent the predicted probability of reporting cognitive decline if every participant were assigned to a given loneliness level, holding other factors constant. We assessed dose–response patterns by comparing marginal probabilities across categories. Pairwise differences were considered statistically significant at p < 0.05.

### Subgroup analyses

To determine whether associations differed by sex, age group, or race/ethnicity, we fitted models including interaction terms between loneliness and each subgroup variable (e.g., i.loneliness##i.sex). Age was categorized into 18–64 and ≥65 years. After estimating models with interactions, we computed subgroup-specific predicted probabilities of SCD for each loneliness category and conducted pairwise comparisons using margins and pwcompare commands. These analyses allowed identification of groups most vulnerable to loneliness-related cognitive decline.

### Sensitivity Analyses

We conducted sensitivity analyses to assess the robustness of our findings. First, we applied inverse probability weighting (IPW) to further reduce potential confounding by baseline characteristics. Propensity scores for the five loneliness categories were estimated using multinomial logistic regression including all covariates in the main model. IPWs were calculated as the inverse of each participant’s predicted probability of being in their observed loneliness category, trimmed at the 1st and 99th percentiles to limit the influence of extreme values, and then multiplied by the BRFSS survey sampling weights to generate final analysis weights. The survey-weighted models were re-estimated using these stabilized IPWs, and effect estimates were consistent with those from the primary analyses.

Second, to address potential bias due to missing data, we performed multiple imputation using chained equations. All exposures, outcomes, and covariates; including auxiliary variables were included in the imputation models. Twenty imputed datasets were generated, and estimates were combined using Rubin’s rules. Results from the multiply imputed analyses were materially unchanged, suggesting that missing data were unlikely to have meaningfully influenced the findings.

### Software

All analyses were conducted using Stata/SE 18.0 (StataCorp LLC, College Station, TX). Code was fully annotated and is available upon reasonable request to facilitate replication.

### Ethics statement

The BRFSS data are publicly available and de-identified; therefore, this secondary analysis did not require institutional review board approval. The study complied with ethical standards for human subjects research and followed guidelines in the Declaration of Helsinki.

## Results

### Baseline Characteristics

Table 1 presents the weighted baseline characteristics of the 86,520 adults included in the analytic sample, stratified by cognitive decline status. Overall, 13,955 participants (16.1%) reported subjective cognitive decline. Several demographics, socioeconomic, and psychosocial characteristics differed significantly between individuals with and without cognitive decline.

**Table 1.** Baseline Sociodemographic Characteristics of Adults by Cognitive Decline Status.

|  | Total Population<br>(N=86, 520) | Cognitive<br>Decline<br>(n=72,565) (No) | Cognitive Decline<br>(n=13,955) (Yes) | Chi-Square | p |
| --- | --- | --- | --- | --- | --- |
| **Age(years)** |  |  |  | 4.484 | 0.112 |
| 18-64yrs. | 39,267 (45.38%) | 33,046 (45.54%) | 6,221 (44.58%) |  |  |
| >64yrs. | 47,253 (54.62%) | 39,519 (54.46%) | 7,734 (55.42%) |  |  |
| **Sex** |  |  |  | 11.168 | <0.01 |
| Male | 38,456 (44.45%) | 32,433 (44.7%) | 6,023 (43.16%) |  |  |
| Female | 48,064 (55.55%) | 40,132 (55.3%) | 7,932 (56.84%) |  |  |
| **Race/Ethnicity** |  |  |  | 33.125 | < 0.01 |
| White | 64,764 (76.4%) | 54,435 (76.5%) | 10,329 (75.68%) |  |  |
| Black | 7,191 (8.48%) | 5,945 (8.4%) | 1,246 (9.13%) |  |  |
| Hispanic | 6,635 (7.83%) | 5,632 (7.9%) | 1,003 (7.35%) |  |  |
| Other | 4,304 (5.08%) | 3,606 (5.1%) | 698 (5.11%) |  |  |
| Multiracial | 1,877 (2.21%) | 1,505 (2.1%) | 372 (2.73%) |  |  |
| **Loneliness Categories** |  |  |  | 4.60E+03 | < 0.01 |
| Never | 38,290 (44.47%) | 34,583 (47.88%) | 3,707 (26.71%) |  |  |
| Always | 2,066 (2.40%) | 1,116 (1.55%) | 950 (6.84%) |  |  |
| Usually | 2,337 (2.71%) | 1,408 (1.95%) | 929 (6.69%) |  |  |
| Sometimes | 17,919 (20.81%) | 13,431 (18.59%) | 4,488 (32.34%) |  |  |
| Rarely | 25,499 (29.61%) | 21,694 (30.03%) | 3,805 (27.42%) |  |  |
| **Education Status** |  |  |  | 359.38 | <0.01 |
| < High School | 25,151 (29.16%) | 20,400 (28.21%) | 4,751 (34.13%) |  |  |
| Some Colleges | 23,418 (27.15%) | 19,343 (26.74%) | 4,075 (29.28%) |  |  |
| College Graduate | 37,675 (43.68%) | 32,582 (45.05%) | 5,093 (36.59%) |  |  |
| **Employment Status** |  |  |  | 918.411 | <0.01 |
| Unemployed | 52,134 (60.63%) | 42,122 (58.42%) | 10,012 (72.14%) |  |  |
| Employed | 33,848 (39.37%) | 29,982 (41.58%) | 3,866 (27.86%) |  |  |
| **Metropolitan Status** |  |  |  | 37.722 | < 0.01 |
| Non-Metropolitan | 24,801 (29.67%) | 20,458 (29.24%) | 4,343 (31.87%) |  |  |
| Metropolitan | 58,796 (70.33%) | 49,510 (70.76%) | 9,286 (68.13%) |  |  |
| **County Type** |  |  |  | 9.243 | <0.01 |
| Rural Counties | 12,370 (14.8%) | 10,238 (14.63%) | 2,132 (15.64%) |  |  |
| Urban Counties | 71,227 (85.2%) | 59,730 (85.37%) | 11,497 (84.36%) |  |  |
| **Marital Status** |  |  |  | 454.128 | <0.01 |
| Married | 47,955 (55.78%) | 41,214 (57.18%) | 6,741 (48.54%) |  |  |
| Single | 7,785 (9.06%) | 6,496 (9.01%) | 1,289 (9.28%) |  |  |
| Divorced | 13,301 (15.47%) | 10,578 (14.68%) | 2,723 (19.61%) |  |  |
| Widowed | 13,555 (15.77%) | 11,137 (15.45%) | 2,418 (17.41%) |  |  |
| Separated | 1,469 (1.71%) | 1,108 (1.54%) | 361 (2.6%) |  |  |
| Member of an unmarried couple? | 1,904 (2.21%) | 1,548 (2.15%) | 356 (2.56%) |  |  |
| **Language** |  |  |  | 46.128 | <0.01 |
| Spanish | 4,280(4.95%) | 3,749 (5.17%) | 531 (3.81%) |  |  |
| English | 82,240 (95.05%) | 68,816 (94.83%) | 13,955 (96.19%) |  |  |

Age distributions were comparable across groups, with adults aged >64 years representing slightly more than half of both those with and without cognitive decline (55.4% vs. 54.5%; p = 0.112). Women constituted a higher proportion of respondents with cognitive decline than those without (56.8% vs. 55.3%; p < 0.01).

Marked differences emerged across racial and ethnic groups (p < 0.01). Although the majority of respondents in both groups were White, Black and multiracial adults represented larger shares of those with cognitive decline. Psychosocial factors also varied substantially, particularly loneliness (p < 0.01). Participants with cognitive decline were considerably more likely to report chronic or frequent loneliness, including “always lonely” (6.8% vs. 1.6%) and “usually lonely” (6.7% vs. 2.0%), whereas respondents without cognitive decline more often endorsed “never lonely.”

Socioeconomic characteristics demonstrated consistent gradients. Individuals with cognitive decline had lower educational attainment, with higher proportions reporting less than a high school education (34.1% vs. 28.2%; p < 0.01) and were more frequently unemployed (72.1% vs. 58.4%; p < 0.01). Geographic characteristics also showed modest but significant variation: respondents with cognitive decline were more likely to reside in non-metropolitan (31.9% vs. 29.2%) or rural counties (15.6% vs. 14.6%; both p < 0.01).

Marital status differed substantially between groups (p < 0.01). Those with cognitive decline had higher proportions who were divorced, widowed, or separated, whereas those without decline were more frequently married. Language preference also varied (p < 0.01), reflecting underlying differences in sample composition.

Collectively, these baseline characteristics highlight pronounced socioeconomic, psychosocial, and demographic differences between adults with and without subjective cognitive decline, underscoring the need for covariate adjustment in subsequent regression analyses.

### Predicted probability of cognitive decline by loneliness category

Loneliness showed a strong dose–response relationship with the likelihood of SCD (Table 1). After adjustment, respondents who reported always feeling lonely had a predicted probability of cognitive decline of 45.7 % (95 % CI, 41.0 – 50.4 %) compared with 38.4 % (95 % CI, 34.4 – 42.5 %) among those who were usually lonely and 24.9 % (95 % CI, 23.6 – 26.1 %) among those who were sometimes lonely. Participants reporting rarely feeling lonely had an estimated probability of 15.0 % (95 % CI, 14.1 – 15.9 %), whereas those who never felt lonely had the lowest probability at 9.9 % (95 % CI, 9.3 – 10.5 %). All pairwise comparisons between loneliness categories were statistically significant (p < 0.001), indicating progressively higher cognitive decline with increasing loneliness.

### Sex differences within loneliness categories

We next tested whether the association between loneliness and cognitive decline differed between women and men. Among participants who always felt lonely, women had a predicted probability of SCD that was 10.7 percentage points higher than men (p = 0.017). In contrast, sex differences were small and not statistically significant among those who were never, rarely, sometimes or usually lonely (differences ranged from –0.9 to –0.2 percentage points; p values > 0.45) (Table 2). These findings suggest that chronic loneliness may disproportionately impact cognitive health in women, whereas occasional loneliness affects men and women similarly.

**Table 2.** Adjusted Predicted Probability of Cognitive Decline by Loneliness Category.

| Loneliness Category | Margin | Std. Err. | t | 95% CI | P-value |
| --- | --- | --- | --- | --- | --- |
| Never | 0.099 | 0.003 | 32.13 | 0.093 – 0.105 | <0.001 |
| Always | 0.457 | 0.024 | 19.09 | 0.410 – 0.504 | <0.001 |
| Usually | 0.384 | 0.021 | 18.58 | 0.344 – 0.425 | <0.001 |
| Sometimes | 0.249 | 0.006 | 38.94 | 0.236 – 0.261 | <0.001 |
| Rarely | 0.150 | 0.005 | 32.92 | 0.141 – 0.159 | <0.001 |

*This table presents the adjusted predicted probabilities of cognitive decline by loneliness category, derived using predictive margins after inverse probability weighting. Loneliness categories include 'Never', 'Rarely', 'Sometimes', 'Usually', and 'Always'. Estimates reflect the adjusted likelihood of self-reported cognitive decline for each category based on BRFSS data. Models were adjusted for age, race/ethnicity, sex, education, employment status, English proficiency, metropolitan status, urbanicity, marital status, and incorporated state and survey year fixed effects. All associations were statistically significant at $P < 0.001$ .*

### Racial and ethnic differences within loneliness categories

We examined racial/ethnic disparities by comparing non-Hispanic Black and Hispanic adults with non-Hispanic White adults within each loneliness category (Table 3). Among respondents who never felt lonely, Black adults had a 1.7-percentage-point lower predicted probability of SCD than White adults (p = 0.036), while the Hispanic–White difference was non-significant (–2.3 percentage points; p = 0.095). Among those who were always or usually lonely, neither Black nor Hispanic participants differed significantly from White participants (p values 0.12 – 0.90). Similarly, within the sometimes and rarely lonely groups, differences between racial/ethnic minorities and White adults were small and not statistically significant. Overall, the magnitude of racial/ethnic disparities was modest compared with the large increases in cognitive decline associated with higher loneliness.

**Table 3.** Sex Differences in the Association Between Loneliness and Cognitive Decline.

| Loneliness Category | Female vs. Male (Difference) | Std. Err. | t | 95% CI | P-value |
| --- | --- | --- | --- | --- | --- |
| Never | -0.009 | 0.0057 | -1.57 | -0.020 to 0.002 | 0.116 |
| Always | 0.107 | 0.0450 | 2.38 | 0.019 to 0.195 | 0.017 |
| Usually | -0.00 | 0.0408 | -0.00 | 0.0800 to 0.0797 | 0.997 |
| Sometimes | -0.002 | 0.0133 | -0.16 | -0.028 to 0.024 | 0.875 |
| Rarely | -0.007 | 0.0087 | -0.75 | -0.024 to 0.010 | 0.452 |

*Table displays the adjusted marginal contrasts comparing females to males in the predicted probability of reporting cognitive decline across loneliness categories. Estimates reflect the sex-specific difference in predicted probability within each loneliness category, adjusted for age, race/ethnicity, education, employment, English proficiency, marital status, metropolitan status, urbanicity, and state and year fixed effects. Positive values indicate higher predicted probability among females relative to males.*

### Age differences within loneliness categories

We explored whether age modified the association between loneliness and cognitive function using an interaction between age group (18–64 years vs >64 years) and loneliness categories (Table 4). Overall, age-related differences in cognitive function were small and generally not statistically significant across most loneliness categories.

**Table 4.**
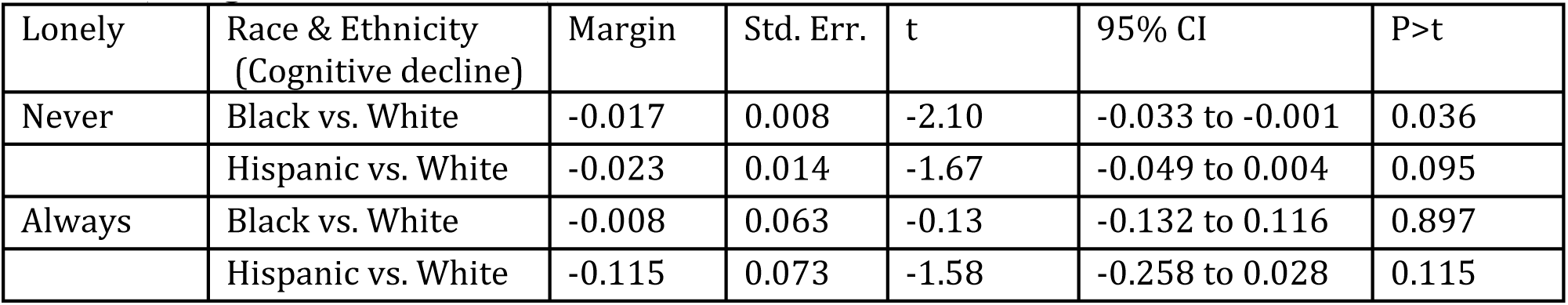
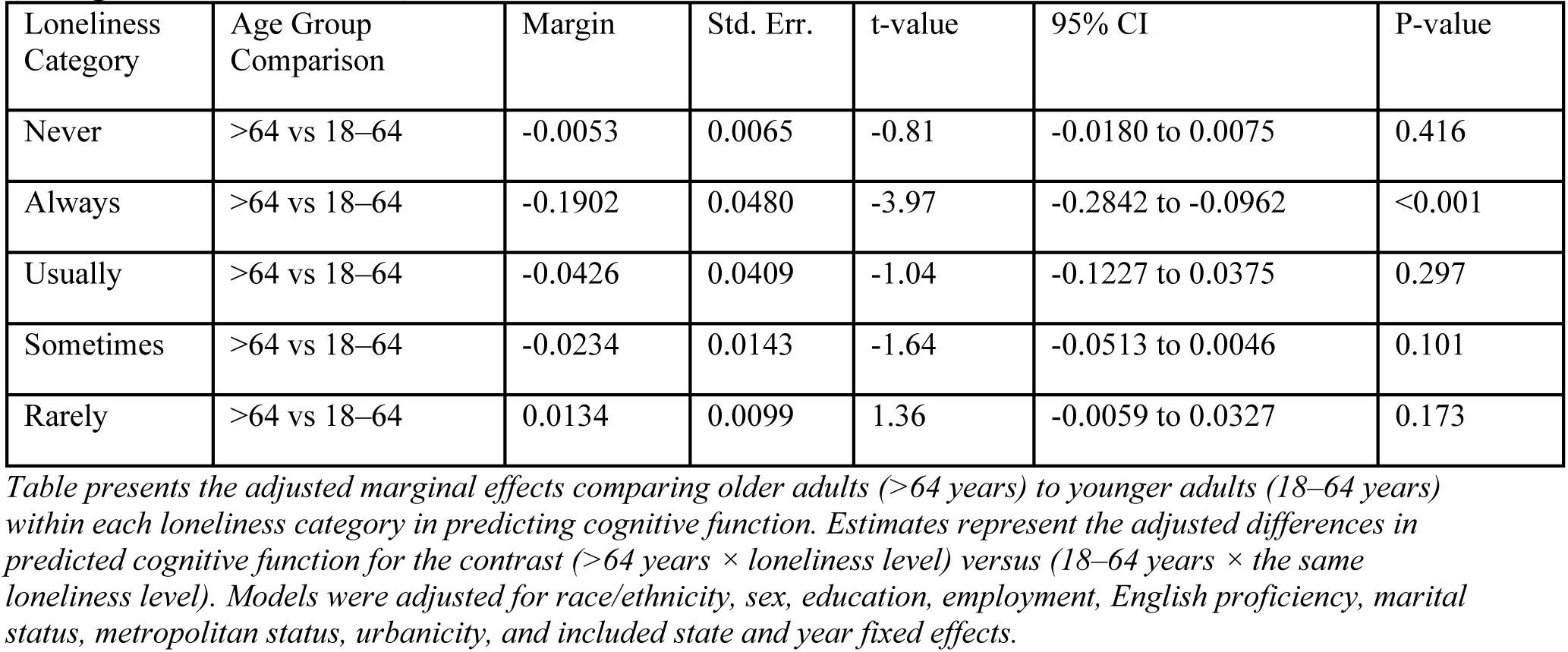
Racial and Ethnic Differences in the Association Between Loneliness and Cognitive Decline (Marginal Effects Model)

Among respondents who were never lonely, older adults (>64 years) did not differ significantly from younger adults (18–64 years) in predicted cognitive function (margin = –0.0053; p = 0.416). Similarly, no meaningful age differences were observed among individuals who reported being sometimes lonely (margin = –0.0234; p = 0.101) or rarely lonely (margin = 0.0134; p = 0.173).

In contrast, a notable divergence emerged among participants who were always lonely. Older adults had significantly lower predicted cognitive function compared with younger adults (margin = –0.1902; p < 0.001), suggesting that the detrimental impact of chronic loneliness may be more pronounced among older adults. Confidence intervals were moderately wide but the effect remained statistically robust.

Among participants who were usually lonely, older adults also showed a lower predicted cognitive function relative to younger adults, but this difference did not reach statistical significance (margin = –0.0426; p = 0.297).

Taken together, these findings indicate that age does not substantially modify the loneliness–cognition relationship when loneliness is mild or intermittent. However, when loneliness is persistent and severe (“always lonely”), older adults appear to experience disproportionately poorer cognitive function compared with younger adults, suggesting heightened vulnerability to the cognitive consequences of chronic loneliness in later life.

## Discussion

This study provides a nationally representative analysis of how self-reported loneliness relates to subjective cognitive decline (SCD) among U.S. adults aged 18 years and older. We observed a clear dose–response pattern: the predicted probability of SCD increased progressively from those who never felt lonely (10 %) to those who rarely and sometimes felt lonely (15 % and 25 %, respectively) and was highest among respondents who usually or always felt lonely (38 % and 46 %). Chronic loneliness was therefore associated with a two- to four-fold greater likelihood of perceived cognitive decline compared with never feeling lonely. These associations persisted after weighing and adjustment for demographic factors. We also found that women who reported persistent loneliness had higher predicted probabilities of cognitive decline than their male counterparts, whereas sex differences were negligible among participants with occasional or no loneliness. Racial/ethnic differences were modest, with a slightly lower risk of cognitive decline among Black adults who never felt lonely compared with White adults and no significant disparities within higher-loneliness categories. Age modestly modified the loneliness–cognition relationship. Among individuals who were rarely or intermittently lonely, cognitive function did not differ meaningfully by age group. However, among those who were chronically lonely (“always lonely”), older adults exhibited significantly lower predicted cognitive function compared with younger adults, suggesting that the cognitive impact of persistent loneliness may be more pronounced in later life.

### Interpretation and implications

The graded association between loneliness and SCD aligns with a broad body of evidence linking social disconnection to cognitive impairment(35–37). Loneliness can provoke chronic activation of stress response systems, leading to dysregulated cortisol secretion and inflammatory pathways that may damage hippocampal and prefrontal brain regions involved in memory and executive function(22, 38, 39). Loneliness may also reduce cognitive stimulation, diminish emotional support, and contribute to depressive symptoms, all of which can undermine cognitive resilience(19, 40, 41). The substantially higher risk of SCD among participants who were always or usually lonely underscores the importance of duration and intensity of loneliness; occasional episodes may be less detrimental, whereas chronic loneliness likely exerts cumulative neurobiological and psychosocial stress. These findings support calls to recognize loneliness as a modifiable risk factor for declined cognition and dementia, alongside other behavioral risk factors identified by the Lancet Commission(19, 42, 43).

Our observation that women experiencing persistent loneliness were more likely than men to report cognitive decline echoes prior research suggesting that loneliness may affect men and women differently(44–46). Some studies have found that men report higher prevalence of loneliness than women, yet women may be more vulnerable to the emotional and physiological consequences of chronic loneliness(47–49). Women often play central roles in maintaining family and social networks; when these networks weaken, the resulting loneliness may carry greater psychological burden(44, 50). Sex differences in neuroendocrine responses to stress, in the prevalence of depression, and in help-seeking behaviors may also contribute (51, 52). In contrast, the lack of sex differences in less intense loneliness categories suggests that occasional loneliness affects men and women similarly(47). Future research should explore gender-specific coping strategies and determine whether tailored interventions are needed.

The modest racial/ethnic differences found in our study contrast with concerns that loneliness may disproportionately impact minority populations(53). Among respondents who never felt lonely, non-Hispanic Black adults had slightly lower predicted probabilities of cognitive decline than White adults. This finding may reflect stronger family and community networks in some Black communities, culturally distinct interpretations of loneliness, or differences in reporting SCD. For Latino adults, the psypost summary of a U.S. cohort study reported that loneliness measured by a three-item scale was actually associated with better cognitive function, suggesting complex cultural differences in how loneliness relates to cognition(54–56). In our data, disparities were small and non-significant among those reporting any degree of loneliness. These results underscore the need to consider cultural context and measurement issues when assessing loneliness and cognitive health.

Age patterns in our analysis were subtle. Across most loneliness categories, including those who were rarely or sometimes lonely, older adults (>64 years) did not differ meaningfully in cognitive function compared with adults aged 18–64 years, suggesting that mild or occasional loneliness may exert similar cognitive effects across the adult lifespan. However, among individuals who were chronically lonely (“always lonely”), older adults exhibited significantly poorer predicted cognitive function than younger adults. This pattern aligns with evidence that persistent loneliness may compound age-related vulnerability through reduced cognitive reserve, heightened neural susceptibility, or preclinical cognitive changes that intensify the cognitive impact of chronic social isolation in later life(19, 37, 38). Although the confidence intervals were moderately wide, the direction and magnitude of the effect suggest that older adults may be particularly sensitive to the neurocognitive consequences of enduring loneliness. Longitudinal studies with larger samples of chronically lonely older adults are needed to better characterize these age-by-loneliness dynamics and clarify underlying mechanisms.

### Policy and public health implications

The strong association between chronic loneliness and cognitive decline has several policy implications. First, public health surveillance systems should continue to monitor loneliness as a key social determinant of cognitive health. Including loneliness measures in national surveys (such as the BRFSS) enables identification of high-risk groups and evaluation of interventions. Second, health systems and community organizations should integrate loneliness screening into routine care for adults, particularly women and mid-life adults. Brief validated tools can help clinicians identify patients who might benefit from referral to social support programs, counselling, or cognitive training. Third, interventions that reduce loneliness – such as social prescribing, group activities, befriending programs, technology-based connections and community engagement initiatives should be scaled up. Evidence from randomized trials indicates that enhancing social support can improve mental health and quality of life; future trials should assess cognitive outcomes. Finally, policies addressing structural drivers of loneliness, including poverty, housing instability, age-friendly environments, and digital inclusion, may have downstream benefits for cognitive health. Given the projected increase in dementia prevalence and the widespread perception of a “loneliness epidemic”, addressing loneliness should be a public health priority.

### Limitations

This study has important limitations. First, the BRFSS data are cross-sectional, precluding causal inference; it is possible that early cognitive decline leads to social withdrawal and loneliness rather than vice versa. Second, both loneliness and cognitive decline were measured by single self-reported items, which may be subject to misclassification and social desirability bias; objective cognitive testing and multi-item loneliness scales would provide more reliable assessments. Third, we lacked information on potentially confounding factors such as depressive symptoms, physical health conditions, sleep quality, socioeconomic status, and social network size, all of which may influence both loneliness and cognition. Fourth, small sample sizes in some strata (e.g., older adults who were always lonely) limited precision and may have produced unstable estimates. Fifth, our analyses focused on subjective cognitive decline rather than clinical diagnoses; however, subjective decline is a known early indicator of objective cognitive impairment. Finally, the BRFSS is restricted to non-institutionalized adults and excludes those living in nursing homes or with severe cognitive impairment, which may underestimate the burden of loneliness and cognitive decline in the broader population.

### Directions for future research

Longitudinal studies are needed to disentangle the temporal ordering of loneliness and cognitive decline and to examine whether reducing loneliness can slow cognitive deterioration. Future research should incorporate comprehensive loneliness instruments that distinguish social isolation, emotional loneliness and duration, as well as objective cognitive assessments and biomarkers of neurodegeneration. Studies should explore potential mediators (e.g., depression, physical activity, sleep, inflammatory markers) and moderators (e.g., socioeconomic status, digital connectivity) of the loneliness–cognition link. Given the heterogeneous findings across racial/ethnic groups, culturally adapted instruments and community-engaged research are critical to understand how loneliness is experienced and reported. Intervention trials targeting loneliness among diverse populations and life stages should measure cognitive outcomes to inform evidence-based policies. Investigating the long-term impact of social disruptions like the COVID-19 pandemic on loneliness and cognitive health is also essential.

## Conclusion

In summary, this study demonstrates a robust dose–response relationship between loneliness and subjective cognitive decline among U.S. adults. Persistent loneliness is associated with markedly higher predicted probabilities of cognitive decline, especially among women and middle-aged adults. Racial/ethnic differences were modest, and age patterns were nuanced, with occasional loneliness in later life conferring substantial risk. These findings reinforce loneliness as an important, modifiable determinant of cognitive health and highlight the urgency of screening for loneliness and implementing targeted social interventions. Addressing the social and structural determinants of loneliness could complement traditional dementia prevention strategies and help preserve cognitive function across the life course.

## Data Availability

Information and data used in this study are available from the corresponding author upon reasonable request.

https://www.cdc.gov/brfss/index.html

---

**Table S5.**
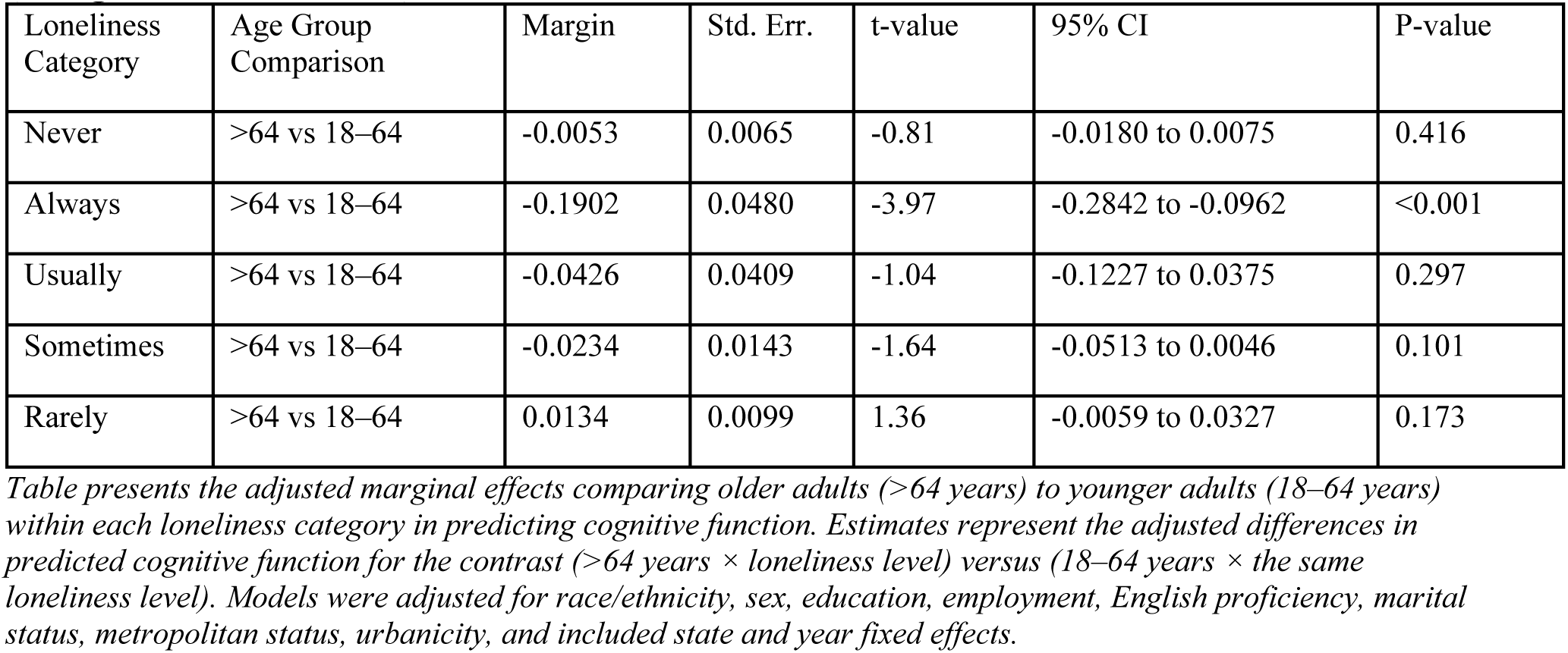
Age Differences in the Association Between Loneliness and Cognitive Decline (Marginal Effects Model)

